# Breakthrough SARS-CoV-2 infections in 620,000 U.S. Veterans, February 1, 2021 to August 13, 2021

**DOI:** 10.1101/2021.10.13.21264966

**Authors:** Barbara A. Cohn, Piera M. Cirillo, Caitlin C. Murphy, Nickilou Y. Krigbaum, Arthur W. Wallace

## Abstract

National data on COVID-19 vaccine breakthrough infections is inadequate but urgently needed to determine U.S. policy during the emergence of the Delta variant. We address this gap by comparing SARS CoV-2 infection by vaccination status from February 1, 2021 to August 13, 2021 in the Veterans Health Administration, covering 2.7% of the U.S. population. Vaccine protection declined by mid-August 2021, decreasing from 91.9% in March to 53.9% (p<0.01, n=619,755). Declines were greatest for the Janssen vaccine followed by Pfizer–BioNTech and Moderna. Patterns of breakthrough infection over time were consistent by age, despite rolling vaccine eligibility, implicating the Delta variant as the primary determinant of infection. Findings support continued efforts to increase vaccination and an immediate, national return to additional layers of protection against infection.

## Background

The messenger RNA vaccines BNT162b2 (Pfizer–BioNTech) and mRNA-1273 (Moderna) and the viral vector vaccine JNJ-78436735 (Janssen) have effectively prevented clinically significant disease caused by severe acute respiratory syndrome coronavirus 2 (SARS-CoV-2) since their rollout in the U.S. in late 2020. Vaccines have also reduced the incidence of asymptomatic infection and associated infectivity (*1*). However, the U.S. has experienced a recent surge in cases of COVID-19, evident in July 2021 and dominated by the B.1.617.2 (Delta) variant (*2*). Although initial reports on vaccine effectiveness, including the six-month follow-up of the Pfizer-BioNTech trial (*3*), suggested sustained protection against infection and hospitalization (*4-6*), breakthrough infections have continued to emerge in vaccine recipients.

This phenomenon has been most comprehensively monitored in Israel, where breakthrough infections, hospitalizations, and deaths have occurred – despite high levels of vaccination in the population (*7*). As a result, Israel authorized boosters of the Pfizer-BioNTech vaccine for adults age ≥ 60 years in July 2021 and extended this authorization to adults age ≥ 50 years on August 13, 2021 (*8*). The U.S. subsequently announced their intention to offer boosters by late September, with the goal to stem the surge in SARS-CoV-2 infections caused by the Delta variant (*9*).

Availability of national data on breakthrough infections in the U.S. has been severely limited. The U.S. Centers for Disease Control and Prevention (CDC) transitioned in May 2021 from monitoring all reported vaccine breakthrough cases to focus on identifying and investigating only hospitalized or fatal cases due to any cause, including causes not related to COVID-19 (*10*). Here, we address this gap and examine infections by vaccination status in 619,755 Veterans during the period February 1, 2021 to August 13, 2021, encompassing the emergence and dominance of the Delta variant in the U.S.

## Materials and Methods

We examined SARS CoV-2 infections in U.S. Veterans age ≥18 years and receiving care in the Veterans Health Administration (VHA), the largest integrated health system in the country. After the U.S. Food and Drug Administration issued an emergency use authorization for the Pfizer-BioNTech vaccine in December 2020, the Department of Veterans Affairs (VA) first provided vaccinations to front-line health care workers and Veterans residing in long-term care facilities in 37 of its medical centers across the U.S.^11^ As vaccine supplies increased, additional Veterans received vaccinations based on age, existing health conditions, and other factors for increased risk of severe illness or death from COVID-19, and as of mid-August 2021, nearly 3.3 million Veterans have been fully vaccinated.

We used the VA Corporate Data Warehouse (CDW) to identify vaccination status (fully vaccinated vs. unvaccinated), vaccine type (Pfizer-BioNTech, Moderna, Janssen), and SARS CoV-2 infections during the period February 1, 2021 to August 13, 2021.We focused on this time period because it encompasses the time when many Veterans became fully vaccinated (i.e., vaccine eligibility extended beyond long-term care facilities) and the recent surge in cases in the U.S. The VA CDW provides discrete, individual-level data, including demographics, administrative claims-based diagnosis and procedure codes, prescriptions, anthropometric measures, and free-text data including procedure notes and pathology reports; data include all 50 states and U.S. territories. Fully vaccinated was defined as two doses of Pfizer-BioNTech or Moderna or one dose of Janssen vaccines, administered at the appropriate intervals. We excluded partial vaccinations and vaccinations that were administered off-label and/or not according to recommendation. SARS-CoV-2 infection was defined as the detection of SARS-CoV-2 on most recent reverse-transcriptase–polymerase-chain-reaction (RT-PCR) assay.

We used Cox proportional hazards models to examine the associations of vaccination status (unvaccinated vs. Pfizer-BioNTech vs. Moderna vs. Janssen) and infection, with vaccination modeled as time-varying. Modeling vaccination as time-varying assigns follow-up time for Veterans before the date of full vaccination (defined as 14 days after receipt of the second dose of Pfizer-BioNTech or Moderna or one dose of Janssen vaccines) as unvaccinated time and time after the date of full vaccination as vaccinated time. We required Veterans to receive a RT-PCR assay to contribute vaccinated and/or unvaccinated follow-up time, such that Veterans who received a recent RT-PCR assay before vaccination contributed unvaccinated time only. Veterans in both groups were followed from February 1, 2021 until their most recent RT-PCR assay or August 13, 2021.

We report hazard ratios (HR) and 95% confidence intervals (CI), adjusted for age and comorbidity. Comorbidity was measured using the Charlson comorbidity index^12^ and a diagnosis of diabetes, chronic obstructive pulmonary disease, bronchitis, acute respiratory failure, chronic lung disease, cardiovascular disease in the two years prior to the RT-PCR assay. We also examined time dependence by including product terms for vaccination status by the log of follow-up time, with p<0.01 indicating statistical significance.

To illustrate findings, we plotted cumulative incidence of infection using Kaplan-Meier estimation to account for censoring, overall and by vaccination status and age group (<50 years, 50-64 years, and ≥65 years). We selected these age groups because they correspond to the phased-in eligibility for vaccination.

All analyses were conducted using SAS Enterprise Guide 7.1 (SAS Institute, Cary, NC). This study was approved by the Institutional Review Board at the University of California San Francisco and the Public Health Institute, as well as the San Francisco VA Research and Development Committee.

## Results

**Table 1** shows the distribution of SARS-CoV-2 infection by demographics and vaccination status for February 1, 2021-August 13, 2021. The percentage of PCR test positivity is higher in Veterans who were unvaccinated (16.6%), female (10.2%), Hispanic (8.9%), American Indian/Alaska Native (9.7%) or Native Hawaiian/Pacific Islander (9.8%), and age <50 years at RT-PCR assay (13.6%).

**Table 1.**
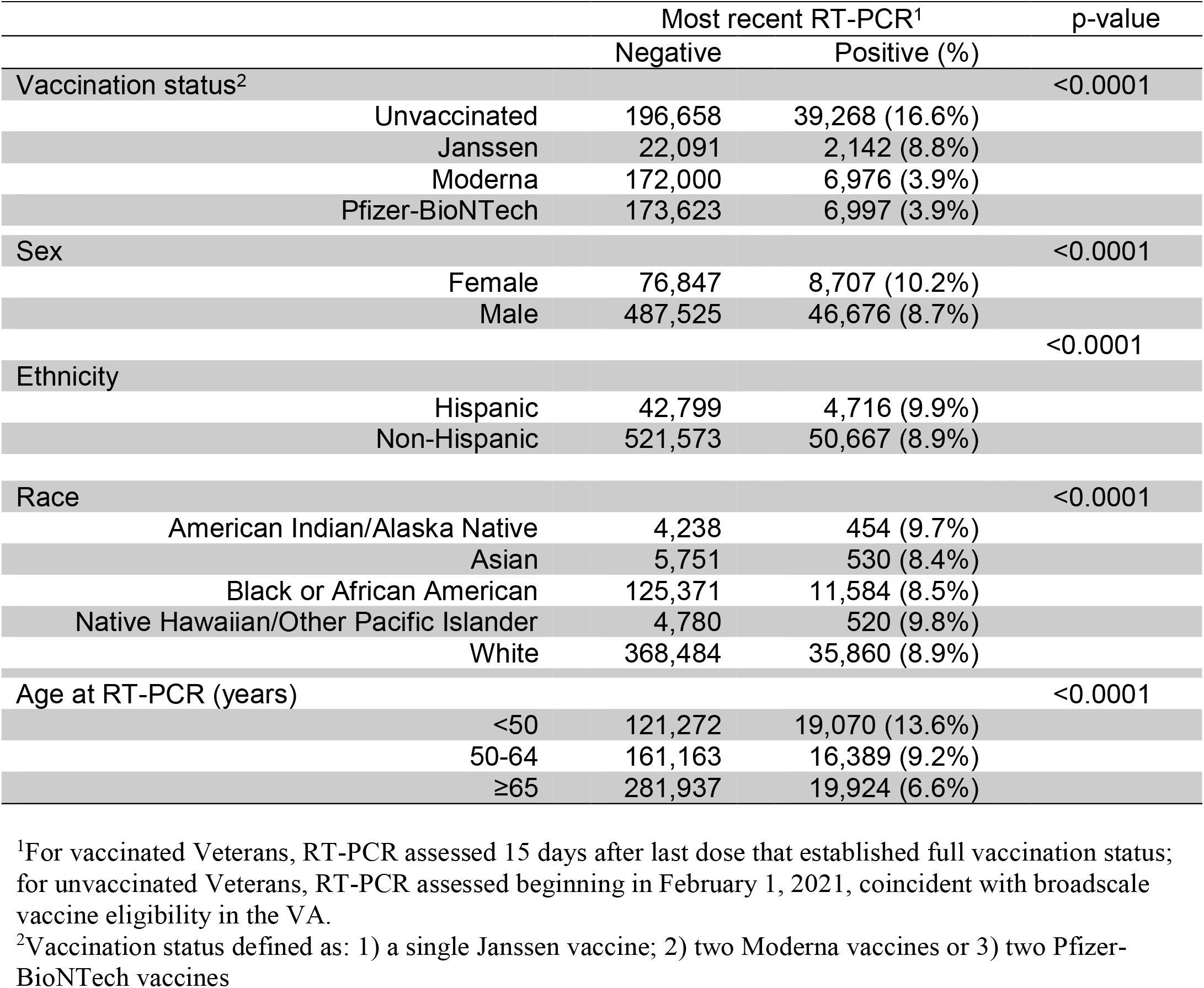
Distribution of SARS-CoV-2 infection by demographics and vaccination status in 619,755 U.S. Veterans, February 1, 2021-August 13, 2021.

For the period February 1, 2021 – August 13, 2021, full vaccination with either Pfizer-BioNTech (aHR 0.16, 95% CI 0.16, 0.17) or Moderna (aHR: 0.13, 95% CI 0.12, 0.13) was associated with lower risk of infection. For the period mid-March 2021 – August 13, 2021, full vaccination with Janssen was also associated with a lower risk of infection (aHR 0.30, 95% CI 0.28, 0.32), owing to the fact that full vaccination with the Janssen vaccine was not possible until March because of timing of authorization. However, these protective associations declined over time (p<0.01 for time dependence, **Table 2**), even after adjusting for age and comorbidity. The proportionate reduction in infection associated with vaccination declined for all vaccine types, with the largest declines for Janssen followed by Pfizer-BioNTech and Moderna (**Figure 1**).Specifically, in March, protection against infection was: 88% (95% CI, 87% to 89%) for Janssen; 92% (95% CI, 92% to 93%) for Moderna; and 91% (95% CI, 91% to 92%) for Pfizer-BioNTech. By August, protection against infection had declined to: 3% (95% CI, -7% to 12%) for Janssen; 64% (95% CI, 62%-66%) for Moderna; and 50% (95% CI, 47% to 52%) for Pfizer-BioNTech.

**Table 2.** Associations of SARS-CoV-2 infection^1^ with vaccination status by month after vaccination^2^,estimated from Cox proportional hazards models^3^ and adjusted for age, race, ethnicity, sex, and comorbidity.

|  | Adjusted hazard ratio | 95% confidence interval |  | p-value |
| --- | --- | --- | --- | --- |
| Janssen vs. unvaccinated<sup>4</sup> |  |  |  |  |
| March | 0.1165 | 0.1025 | 0.1325 | <0.0001 |
| April | 0.1780 | 0.1613 | 0.1965 | <0.0001 |
| May | 0.2719 | 0.2524 | 0.2930 | <0.0001 |
| June | 0.4153 | 0.3896 | 0.4427 | <0.0001 |
| July | 0.6343 | 0.5898 | 0.6822 | <0.0001 |
| August | 0.9689 | 0.8804 | 1.0662 | 0.5172 |
| Moderna vs unvaccinated<sup>4</sup> |  |  |  |  |
| March | 0.0730 | 0.0694 | 0.0767 | <0.0001 |
| April | 0.1003 | 0.0964 | 0.1043 | <0.0001 |
| May | 0.1378 | 0.1333 | 0.1425 | <0.0001 |
| June | 0.1894 | 0.183 | 0.196 | <0.0001 |
| July | 0.2603 | 0.2497 | 0.2714 | <0.0001 |
| August | 0.3577 | 0.3392 | 0.3772 | <0.0001 |
| Pfizer-BioNTech vs. unvaccinated<sup>4</sup> |  |  |  |  |
| March | 0.0855 | 0.0816 | 0.0896 | <0.0001 |
| April | 0.1219 | 0.1176 | 0.1265 | <0.0001 |
| May | 0.1739 | 0.1687 | 0.1792 | <0.0001 |
| June | 0.2480 | 0.2405 | 0.2557 | <0.0001 |
| July | 0.3536 | 0.3408 | 0.3670 | <0.0001 |
| August | 0.5043 | 0.4811 | 0.5287 | <0.0001 |

<sup>1</sup> Adjusted hazard ratio < 1.0 indicates lower risk of infection for vaccine, shown compared to unvaccinated. For vaccinated Veterans, infection is assessed 15 days after the last vaccine that established full vaccination status. For unvaccinated Veterans, infection is assessed beginning in February 1, 2021, coincident with broadscale vaccine eligibility in the VA.
<sup>2</sup>Time dependence was tested in Cox proportional hazards models by including product terms for vaccination status (Janssen, Moderna, Pfizer-BioNTech, unvaccinated) by log(time):Janssen\*log(time), Moderna\*log(time), Pfizer-BioNTech\*log(time); and adjusted for age, sex, race, ethnicity, sex, and comorbidity (Charlson Comorbidity score, overweight, Type II diabetes, Chronic obstructive pulmonary disease, Bronchitis, Acute respiratory failure and Chronic lung disease). Significance levels for all product terms were p<0.0001
<sup>3</sup>Vaccination status is modeled as time-varying, assigning follow-up time for Veterans before the date of full vaccination as unvaccinated time and time after the date of full vaccination as vaccinated time; vaccination is defined as: 1) a single Janssen vaccine; 2) two Moderna vaccines; or 3) two Pfizer-BioNTech vaccines.
<sup>4</sup>Associations at each month were estimated from contrasts using product terms for vaccination status by time in Cox proportional hazards models, including indicator terms for vaccination status (Janssen, Moderna Pfizer-BioNTech) product terms, and age, sex, race, ethnicity, sex, and comorbidity (Charlson Comorbidity score, overweight, Type II diabetes, Chronic obstructive pulmonary disease, Bronchitis, Acute respiratory failure and Chronic lung disease).

**Figure 1.**
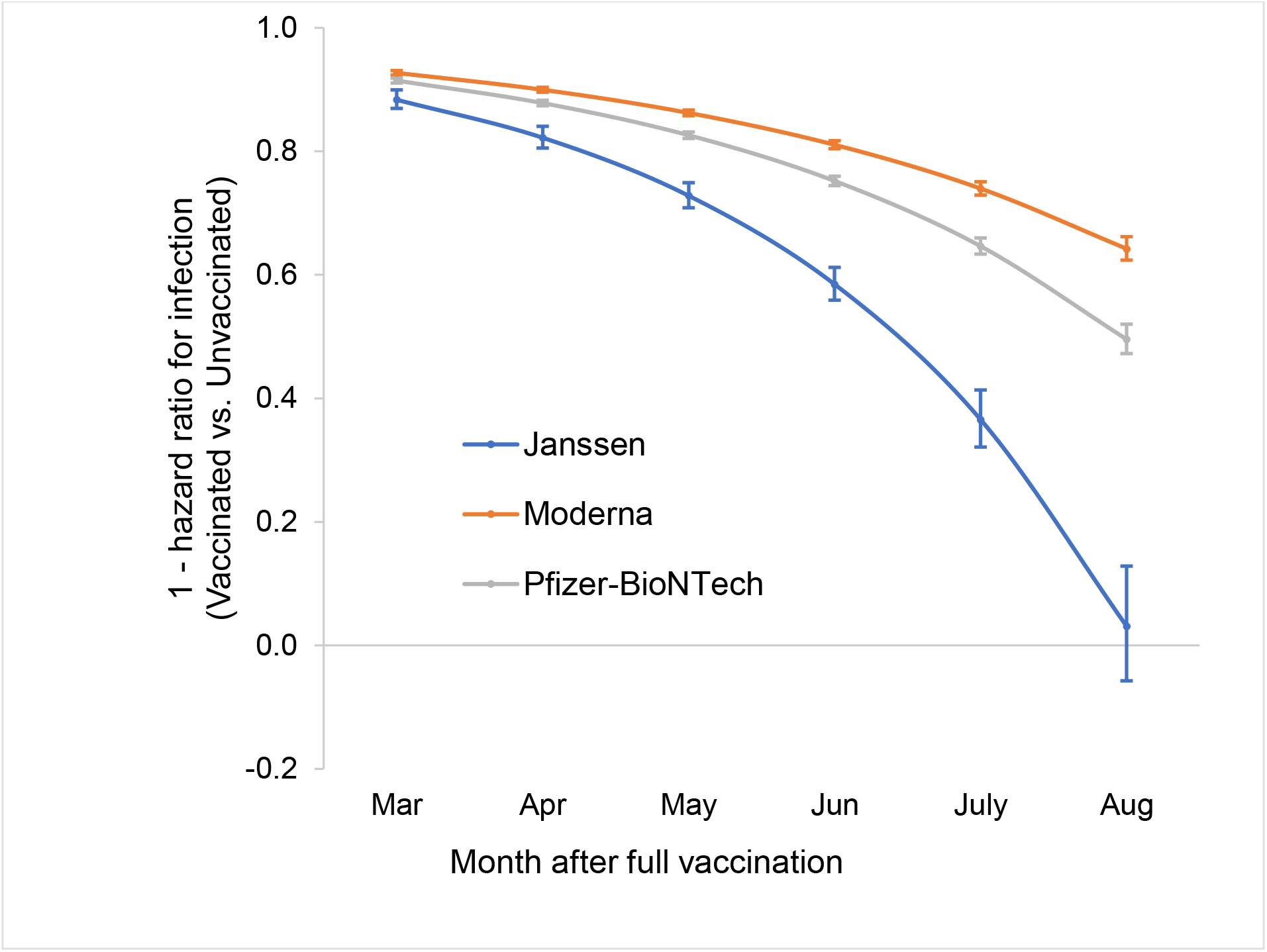
Time dependent vaccine protection against SARS-CoV-2 infection as estimated from Cox proportional hazards models, adjusted for age, race, ethnicity, sex and comorbidity Associations are presented as 1 – hazard ratios and 95% confidence intervals. Associations for each month were estimated from contrasts using product terms for vaccination status by time to most recent RT-PCR.

As shown in **Figure 2**, cumulative incidence of infection accelerated in both unvaccinated and fully vaccinated Veterans beginning in July 2021 and through mid-August 2021, consistent with the time dependence observed in the Cox proportional hazards models. This pattern was similar across age groups; risk of infection was highest for unvaccinated Veterans. Veterans who were fully vaccinated with the Moderna vaccine had the lowest risk of infection, followed closely by those who received the Pfizer-BioNTech vaccine, then those who received the Janssen vaccine.

**Figure 2.**
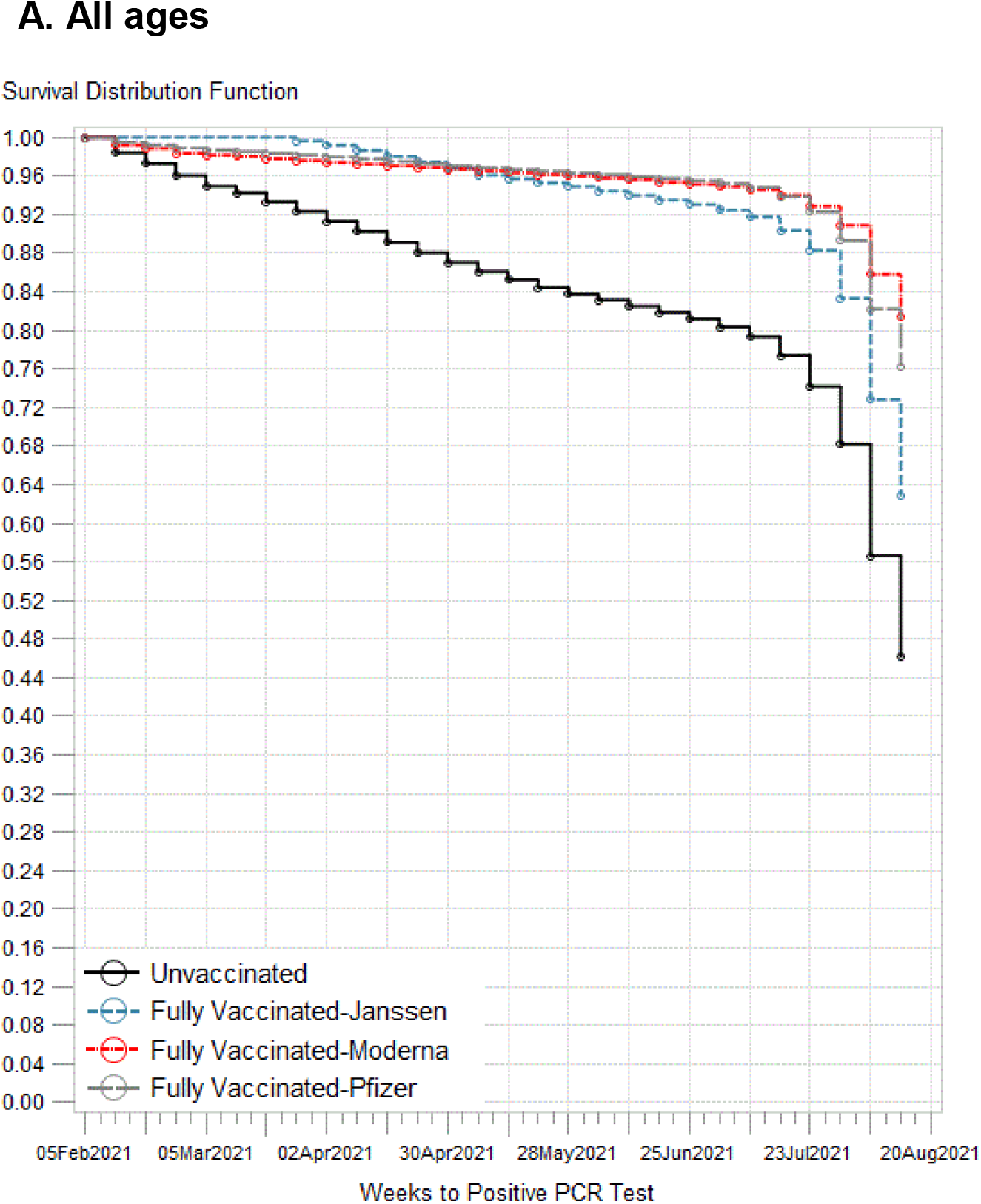
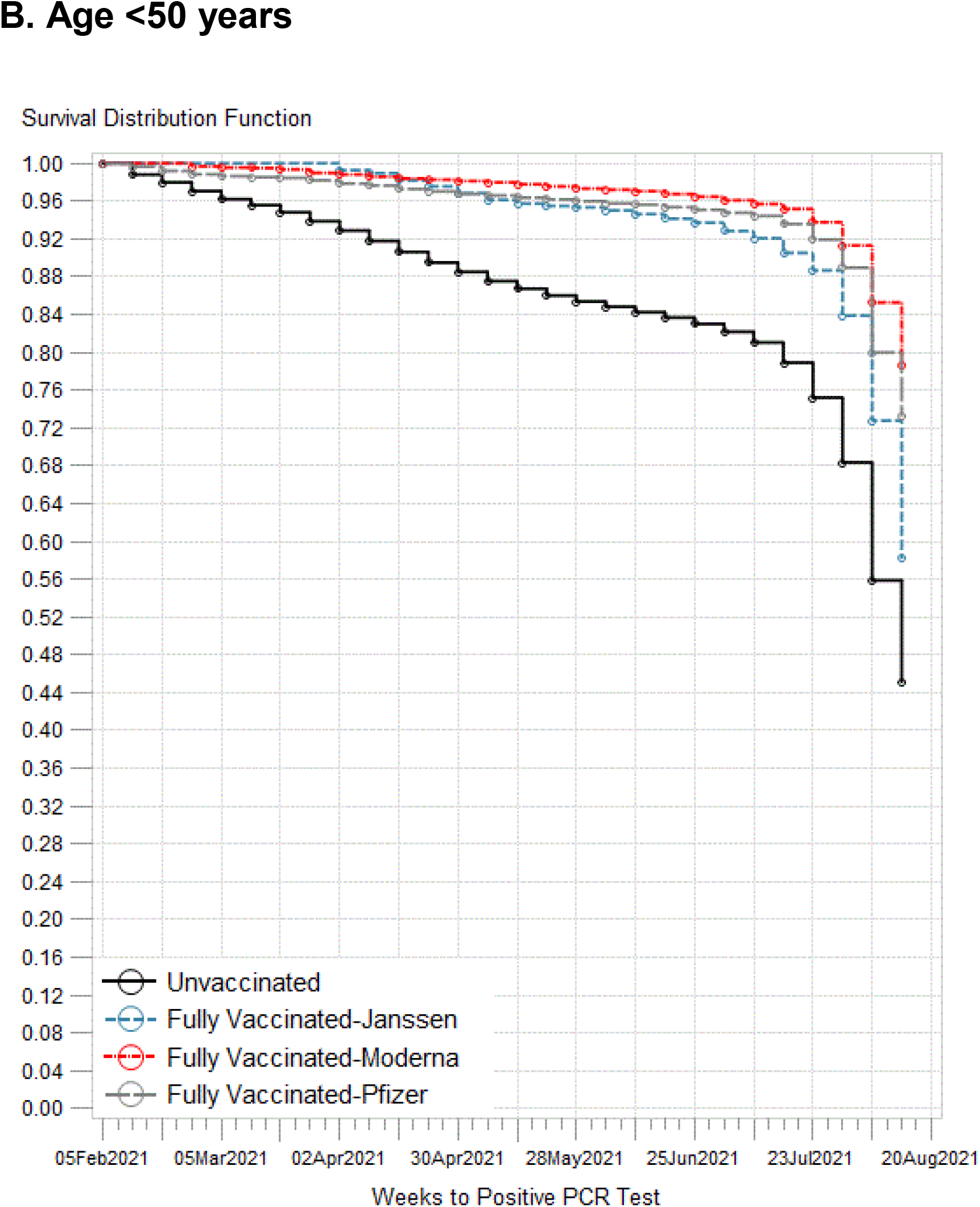
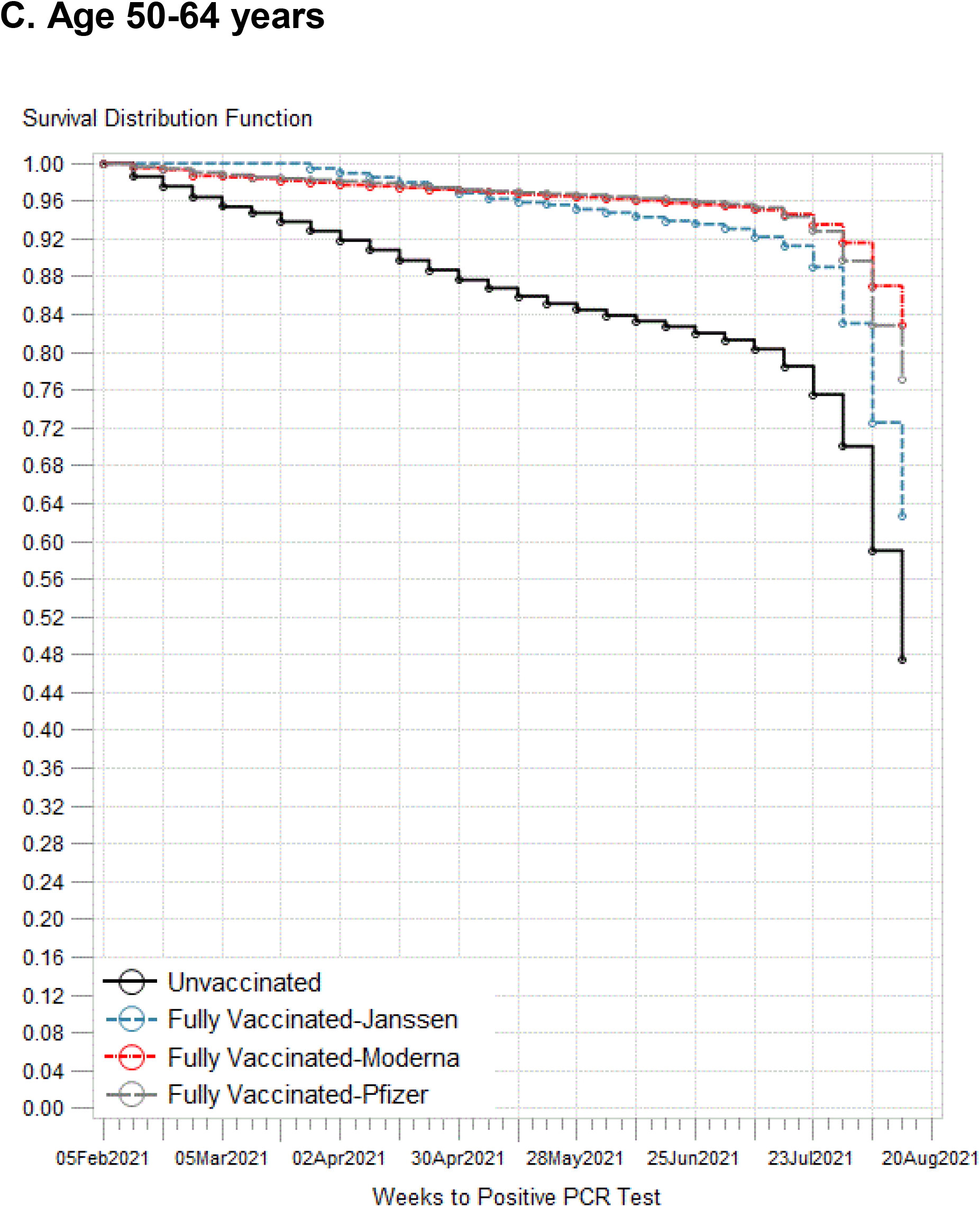
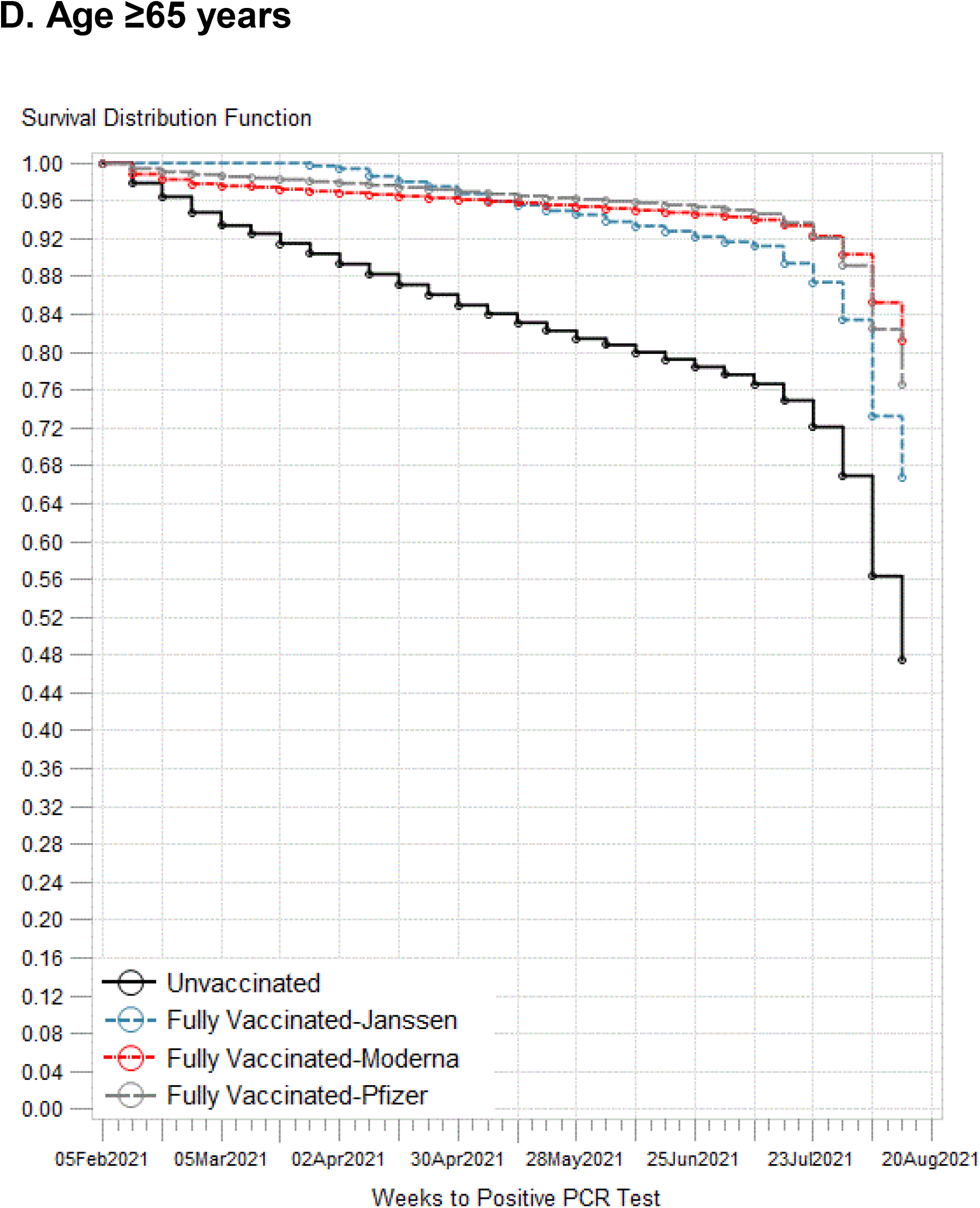
Kaplan-Meier curves illustrating risk of SARS-CoV-2 infection by vaccination status and age: A) all ages; B) age <50 years, C) age 50-64 years; D) age ≥65 years The survival function estimates time to infection detected by most recent RT-PRC.

## Discussion

Benefits of vaccination in reducing risk of SARS-CoV-2 infection are clearly supported by this study of nearly 620,000 U.S. Veterans. However, the protective association between vaccination and infection declined as incidence of infection increased from July 2021 to mid-August 2021. Risk of infection increased in both unvaccinated and vaccinated Veterans, coincident with the emergence and dominance of the Delta variant in the U.S. (*6*). Our analysis of infection by vaccine type, including the Pfizer-BioNTech, Moderna, and Janssen vaccines, suggests waning vaccine protection against infection over time, particularly for the Janssen vaccine. These results demonstrate an urgent need to reinstate multiple layers of protection against infection, such as masking and physical distancing, while also bolstering current efforts to increase vaccination.

Patterns of breakthrough SARS-CoV-2 infection among vaccinated Veterans show a worrisome temporal trend, overlapping with the emergence of Delta as the dominant variant in July 2021. Increasing risk of infection was not explained by age or comorbidity, implicating increased infectivity of the Delta variant vs. waning immunity as the primary determinant of infection. The Delta variant is more infectious than other variants, likely secondary to increased viral load (*2*). In our analysis, the oldest age group (≥65 years) had a similar pattern of breakthrough infections over time compared to the younger age group, despite becoming fully vaccinated 3-4 months earlier, on average. The proportional reduction in vaccine effectiveness against infection across all age groups underscores the critical importance of a layered approach to protection, while enhancing continued efforts to increase vaccination among the 90 million Americans who remain unvaccinated.

Although follow-up of the Pfizer-BioNTech trial demonstrated sustained vaccine protection against infection (91%) (*3*), our results suggest vaccines are less effective in preventing infections with the more recent Delta variant. It is not yet clear whether an additional dose of the same vaccine will confer additional protection against Delta or other variants. On the other hand, Israel’s early experience with boosters suggests a benefit; the number of cases reported in vaccinated persons began to decline in mid-August 2021, within weeks of providing boosters for adults age ≥ 60 years. It is also possible that this decline coincided with behavior change or reinstatement of additional public health interventions.

An important strength of our study is the use of large-scale, national VA data, covering 2.7% of the U.S. population and collected in real time. After it transitioned to focus only on breakthrough hospitalizations and deaths, the CDC created a database for state health departments to enter information on breakthrough infections; however, reporting is not mandatory, and as a consequence, information on risk of breakthrough infections is collected haphazardly across U.S. The VA CDW was essential to our timely analysis of breakthrough infection through mid-August 2021, and moving forward, these data may be used as tool to comprehensively monitor vaccine effectiveness as the Delta variant continues to spread and other variants are likely to emerge.

Our results should be interpreted in the context of limitations. We required a recent RT-PCR assay to be included in the analysis, and there may be differences in testing intervals and frequency by vaccination status. It is also likely that requiring a recent RT-PCR assay does not capture asymptomatic infections, during which testing may not be obtained. Our sample has proportionately fewer women, although a large number are still included. We did not have information on genotyping of infections to determine the proportion caused by the Delta variant. Finally, there is a delay between infection and hospitalization and death that is not captured by the urgency of this report.

In summary, although vaccination remains protective against SARS-CoV-2 infection, this protection is waning as the Delta variant has emerged in the U.S. It is not yet clear whether reductions in vaccine protection against infection will translate into similar reductions in protection against hospitalization and death. COVID-19 vaccines remain the most important tool to prevent infection, severe illness, and death, but vaccines should be accompanied by additional measures, including masking, hand washing, physical distancing, and other public health interventions, in the face of increased risk of infection due to the Delta variant.

## Data Availability

The data that support the findings of this study are available from the Department of Veterans Affairs (VA). Data are made freely available to researchers behind the VA firewall with an approved study protocol. More information is available at https://www.virec.research.va.gov or by contacting the VA Information Resource Center at.

## Acknowledgments

We acknowledge the invaluable efforts of the Veterans Affairs data architects, managers, and clinicians who assembled the Centralized Interactive Phenomics Resource (CIPHER), rapidly compiling a library of numerous COVID-19-related phenotypes that are the basis for this research. We deeply appreciate the steady service and support of the VA Informatics and Computing Infrastructure (VINCI) staff. Without the efforts of these teams, this study would not have been possible. We are grateful for the Veterans who have so selflessly served their country. The views expressed in this article are those of the authors and do not necessarily reflect the position or policy of the Department of Veterans Affairs or the United States government.

## Funding

Mercatus Center at George Mason University (Fast Grants #2207)

University of California Office of the President (Emergency COVID-19 Research Seed Funding R00RG3118)

## Author contributions

Conceptualization: BAC, PMC

Methodology: PMC, CCM, BAC

Statistical analysis: PMC

Funding acquisition: AWW, PMC, NYK

Data interpretation: all authors

Writing - original draft: CCM, PMC, BAC

Writing - review & editing: all authors

## Competing interests

CCM reports consulting for Freenome. AWW reports consulting for ECOM Medical, Obelab, Sensifree, and Shifamed. BAC, PMC, and NYK declare that they have no competing interests.

## Notes

### Author Declarations

This study was approved by the Institutional Review Board at the University of California San Francisco and the Public Health Institute, as well as the San Francisco VA Research and Development Committee.

